# Clinical features of UK Biobank subjects carrying protein truncating variants in genes implicated in schizophrenia pathogenesis

**DOI:** 10.1101/2021.12.07.21267402

**Authors:** David Curtis

**Affiliations:** UCL Genetics Institute, University College London; Centre for Psychiatry, Queen Mary University of London

## Abstract

The SCHEMA consortium has identified ten genes in which protein truncating variants (PTVs) confer substantial risk of schizophrenia. Exome-sequenced participants in the UK Biobank who carried PTVs in these genes were studied to determine to what extent they demonstrated features of schizophrenia or had neuropsychiatric impairment. Following automated quality control and visual inspection of reads, 251 subjects were identified as having well-supported PTVs in one of these genes. The frequency of PTVs in *CACNA1G* was higher than had been observed in SCHEMA cases, casting doubt on its role in schizophrenia pathogenesis, but otherwise rates were similar to those observed in SCHEMA controls. Numbers were too small to allow formal statistical analysis but in general carriers of PTVs did not appear to have high rates of psychiatric illness or reduced educational or occupational functioning. One subject with a PTV in *SETD1A* had a diagnosis of schizophrenia. one with a PTV in *HERC1* had psychotic depression and two subjects seemed to have developmental disorders, one with a PTV in *GRIN2A* and one with a PTV in *RBCC1*. There seemed to be somewhat increased rates of affective disorders among carriers of PTVs in *HERC1* and *RB1CC1*. Carriers of PTVs did not appear to have subclinical manifestations of schizophrenia. Although PTVs in these genes can substantially increase schizophrenia risk, their effect seems to be dichotomous and most carriers appear psychiatrically well.

This research has been conducted using the UK Biobank Resource.

## Introduction

It is now well-established that copy number variants (CNVs) at a number of specific locations can substantially increase risk of developing schizophrenia as well as other neurodevelopmental disorders (Rees *et al*., 2014). Although they have large effects, these CNVs are incompletely penetrant, meaning that not everybody who carries one goes on to develop a neurodevelopmental phenotype. However a recent analysis of the UK Biobank sample has illustrated that even apparently unaffected carriers of 12 CNVs implicated in schizophrenia pathogenesis had on average reduced educational and occupational functioning and reduced performance on cognitive tests (Kendall *et al*., 2019).

Likewise, a study of the same UK Biobank sample showed that, even without a diagnosis of mental illness, subjects with a higher polygenic risk score for schizophrenia were more likely to report experiencing psychotic symptoms (Legge *et al*., 2019). Among patients with schizophrenia, a higher polygenic risk score is associated with greater severity of illness (Jonas *et al*., 2019). These findings suggest that common variants may tend to have a quantitative effect on modifying the frequency and severity of psychotic symptoms for people both with and without a diagnosis of schizophrenia.

The SCHEMA study of exome-sequenced samples of 24,248 cases and 97,322 controls has now implicated ultra-rare coding variants in ten genes as conferring substantial risk for schizophrenia (Singh and on behalf of the Schizophrenia Exome Meta-Analysis (SCHEMA) Consortium, 2022). For these ten genes, exome-wide statistically significant results were obtained from a metanalysis of case-control data and de novo variants identified in trios. Protein truncating variants (PTVs) and damaging missense variants were associated with substantial effects on risk, with odds ratios ranging from 3 to 50. As well as there being an excess of PTVs in cases compared with controls, in the trios *de novo* PTVs in schizophrenia probands were also observed for all these genes apart from *CACNA1G, GRIN2A* and *HERC1*. PTVs in these genes were rare, being observed a total of 127 times among cases and 78 times among controls. For all genes except *GRIA3* a PTV was observed in at least one control, suggesting incomplete penetrance, although it must be borne in mind that control exomes were derived from a variety of cohorts including ExAC and were not all screened for an absence of psychiatric illness.

The UK Biobank consists of a sample of 500,000 volunteers from whom a large amount of phenotypic data has been obtained. Sequence data has been provided for 200,632 subjects who have undergone exome-sequencing, quality control and genotyping by the UK Biobank Exome Sequencing Consortium using the GRCh38 assembly with coverage 20X at 95.6% of sites on average (Szustakowski *et al*., 2020). We investigated this sample to see whether any subjects carrying PTVs in the ten implicated genes demonstrated evidence for indicators of neuropsychiatric problems, including impaired educational or occupational performance.

## Methods

UK Biobank had obtained ethics approval from the North West Multi-centre Research Ethics Committee which covers the UK (approval number: 11/NW/0382) and had obtained informed consent from all participants. The UK Biobank approved an application for use of the data (ID 51119) and ethics approval for the analyses was obtained from the UCL Research Ethics Committee (11527/001). All variants were annotated using variant effect predictor (VEP) (McLaren *et al*., 2016). GENEVARASSOC and SCOREASSOC were used to identify subjects carrying variants annotated as PTVs, consisting of stop, frameshift and essential splice site variants.

For all variants annotated as PTVs, CRAM files were downloaded and calls were rejected if the read depth was less than 20, if the allele balance did not exceed 0.25 or if Fisher’s exact test for distributions of alleles across strands was significant at p<0.01. Additionally, snapshots of the reads produced by the Integrative Genomics Viewer (IGV) were inspected to check that the genotype calls appeared to be well supported (Robinson *et al*., 2017). For any that appeared doubtful, for example where another nucleotide differed from the reference potentially within the same codon, more detailed examination was performed using IGV interactively.

For subjects carrying a well-supported PTV in one of these genes, R was used to extract fields containing clinical information regarding diagnosis, treatment and responses to questions about mental health, along with information reflecting intellectual and occupational functioning, as had been done in a previous study of subjects carrying a PTV in *HIRA* (R Core Team, 2014; Curtis, 2021). Within the UK Biobank dataset, different fields are completed for different subjects and attention was focussed on those fields which were most frequently completed, including fields relating to depression and anxiety, fluid intelligence, reported diagnoses, prescribed medication, educational attainment and employment status. For some of these measures, information was summarised on a per gene basis. It was noted that a large number of subjects carried a PTV in *CACNA1G*, casting doubt on its role as a susceptibility gene, but for all other genes all extracted fields were inspected on an individual basis in order to identify subjects who might have intellectual or occupational impairment or might suffer neuropsychiatric problems. The full list of fields used to provide this information is shown in Supplementary Table 1.

## Results

Across all ten genes a total of 290 variants were initially annotated as PTVs by VEP. Of these, 26 failed the automated QC criteria, of which 22 had also been flagged as possibly problematic by visual inspection of the IGV snapshots. An additional 26 passed the QC but the IGV snapshots appeared possibly problematic for one reason or another and they were subjected to more detailed examination. Of these, there were 6 which were annotated by VEP as “stop_gained” on the basis of a single nucleotide change but in fact another nucleotide in the same codon also differed from the reference, meaning that the overall effect was to produce a non-synonymous amino acid substitution, not a stop codon. An additional 7 calls were judged to be possibly suspect for a number of reasons, such as a lack of supporting reads from one or other strand, and these were also rejected. Supplementary Table 2 provides a list of calls with possibly problematic IGV snapshots along with their read counts and reasons why they were accepted or rejected. These processes resulted in a total of 251 subjects judged to have well-supported PTVs in one of the ten genes.

Table 1 shows summary findings for these subjects on a per gene basis. As well as the raw counts of the numbers of subjects carrying PTVs, the table also depicts this as a prevalence per 100,000 and also for comparison the prevalence in controls and cases in the SCHEMA samples. This makes it clear that for almost all of the genes the prevalence of PTVs in the UK Biobank sample is very similar to that observed in SCHEMA controls (Singh and on behalf of the Schizophrenia Exome Meta-Analysis (SCHEMA) Consortium, 2022). The obvious clear exception is *CACNA1G*, for which we observe a prevalence of 45.9 per 100,000, much higher than the 13.4 observed in SCHEMA controls and even slightly higher than the 41.2 observed in SCHEMA cases. Although a higher prevalence of PTVs might in theory be observed with differences in sequencing technologies producing more thorough coverage or more false positives an alternative explanation is that impaired functioning of *CACNA1G* is not, after all, a risk factor for schizophrenia. In the SCHEMA study the results for *CACNA1G* only just reached criteria for exome-wide statistical significance and PTVs in it were associated with a lower OR than for the other nine genes. In addition, it was one of only three of the implicated genes for which no *de novo* PTV was observed in the trio samples. With these uncertainties in mind, for the present study it was decided not to carry out further detailed investigation of the subjects carrying PTVs in *CACNA1G*. For *RB1CC1*, we observe a prevalence of 12.5 per 100,000, which is three times the prevalence of 4.1 observed in SCHEMA controls. However this is still much lower than the prevalence of 37.1 in SCHEMA cases and so does not cast doubt on the claim that impaired function of *RB1CC1* substantially increases schizophrenia risk.

**Table 1.** Table summarising key phenotypic characteristics of subjects carrying protein-truncating variants (PTVs) in ten genes implicated in schizophrenia risk. For quantitative variables, the mean and SD in the whole UK Biobank sample are provided for reference and for categorical variables the proportions of UK Biobank subjects falling into each category are shown.

|  | Mean (SD) or proportion giving response in whole UK Biobank sample | Gene |  |  |  |  |  |  |  |  |  |
| --- | --- | --- | --- | --- | --- | --- | --- | --- | --- | --- | --- |
|  |  | *SETD1A* | *CUL1* | *XPO7* | *TRIO* | *CACNA1G* | *SP4* | *GRIA3* | *GRIN2A* | *HERC1* | *RB1CC1* |
| **Number of PTV carriers in UK Biobank** |  | 11 | 2 | 1 | 34 | 92 | 6 | 2 | 8 | 70 | 25 |
| **Prevalence expressed as PTV carriers per 100,000** |  |  |  |  |  |  |  |  |  |  |  |
| UK Biobank |  | 5.5 | 1.0 | 0.5 | 16.9 | 45.9 | 3.0 | 1.0 | 4.0 | 34.9 | 12.5 |
| SCHEMA Cases |  | 61.9 | 33.0 | 49.5 | 74.2 | 41.2 | 53.6 | 20.6 | 37.1 | 115.5 | 37.1 |
| SCHEMA Controls |  | 3.1 | 1.0 | 1.0 | 16.4 | 13.4 | 6.2 | 0.0 | 2.1 | 32.9 | 4.1 |
| **Body mass index (BMI)** | 27.3 (4.77) | 28.5 | 28.7 | 29.0 | 26.2 | 27.9 | 28.8 | 27.6 | 31.0 | 27.9 | 29.7 |
| **Age completed full time education** | 16.7 (2.34) | 19.2 | 15.0 | 18.0 | 17.3 | 17.2 | 16.8 | 17.5 | 15.7 | 16.5 | 17.0 |
| **Qualifications** |  |  |  |  |  |  |  |  |  |  |  |
| College or University degree | 0.18 | 6 | 1 | 0 | 6 | 31 | 1 | 0 | 1 | 19 | 4 |
| A levels/AS levels or equivalent | 0.14 | 0 | 0 | 0 | 6 | 4 | 1 | 0 | 1 | 6 | 3 |
| O levels/GCSEs or equivalent | 0.25 | 3 | 0 | 1 | 5 | 20 | 1 | 1 | 1 | 21 | 6 |
| CSEs or equivalent | 0.07 | 0 | 0 | 0 | 3 | 6 | 1 | 1 | 0 | 10 | 3 |
| NVQ or HND or HNC or equivalent | 0.10 | 1 | 0 | 0 | 2 | 3 | 1 | 0 | 0 | 5 | 0 |
| Other professional qualifications<br>eg: nursing, teaching | 0.15 | 1 | 0 | 0 | 2 | 8 | 0 | 0 | 0 | 0 | 0 |
| None of the above | 0.09 | 0 | 1 | 0 | 8 | 20 | 1 | 0 | 4 | 9 | 9 |
| Prefer not to answer | 0.01 | 0 | 0 | 0 | 1 | 0 | 0 | 0 | 1 | 0 | 0 |
| **Current employment status** |  |  |  |  |  |  |  |  |  |  |  |
| In paid employment or self-employed | 0.52 | 7 | 0 | 0 | 12 | 54 | 3 | 2 | 5 | 35 | 10 |
| Retired | 0.32 | 2 | 2 | 1 | 17 | 28 | 2 | 0 | 3 | 24 | 10 |
| Looking after home and/or family | 0.05 | 1 | 0 | 0 | 2 | 4 | 0 | 0 | 0 | 1 | 2 |
| Unable to work because of sickness<br>or disability | 0.04 | 1 | 0 | 0 | 1 | 2 | 0 | 0 | 0 | 6 | 2 |
| Unemployed | 0.02 | 0 | 0 | 0 | 1 | 2 | 0 | 0 | 0 | 2 | 1 |
| Doing unpaid or voluntary work | 0.03 | 0 | 0 | 0 | 0 | 1 | 0 | 0 | 0 | 1 | 0 |
| Full or part-time student | 0.01 | 0 | 0 | 0 | 0 | 1 | 0 | 0 | 0 | 0 | 0 |
| None of the above | 0.01 | 0 | 0 | 0 | 0 | 0 | 0 | 0 | 0 | 1 | 0 |
| Prefer not to answer | 0.00 | 0 | 0 | 0 | 0 | 0 | 1 | 0 | 0 | 0 | 0 |
| **Overall health rating** |  |  |  |  |  |  |  |  |  |  |  |
| Excellent | 0.16 | 1 | 0 | 0 | 6 | 14 | 0 | 0 | 2 | 5 | 6 |
| Good | 0.58 | 6 | 1 | 1 | 19 | 54 | 4 | 1 | 2 | 38 | 10 |
| Fair | 0.21 | 4 | 1 | 0 | 7 | 19 | 2 | 0 | 4 | 23 | 6 |
| Poor | 0.05 | 0 | 0 | 0 | 1 | 4 | 0 | 0 | 0 | 4 | 2 |
| Do not know | 0.00 | 0 | 0 | 0 | 0 | 0 | 0 | 0 | 0 | 0 | 1 |
| Prefer not to answer | 0.00 | 0 | 0 | 0 | 0 | 1 | 0 | 1 | 0 | 0 | 0 |
| **Seen doctor (GP) for nerves, anxiety, tension or depression** |  |  |  |  |  |  |  |  |  |  |  |
| No | 0.65 | 5 | 1 | 1 | 20 | 58 | 4 | 1 | 6 | 32 | 11 |
| Yes | 0.34 | 6 | 1 | 0 | 13 | 32 | 2 | 0 | 2 | 37 | 14 |
| Do not know | 0.01 | 0 | 0 | 0 | 0 | 1 | 0 | 0 | 0 | 0 | 0 |
| Prefer not to answer | 0.00 | 0 | 0 | 0 | 0 | 1 | 0 | 1 | 0 | 1 | 0 |
| **Seen a psychiatrist for nerves, anxiety, tension or depression** |  |  |  |  |  |  |  |  |  |  |  |
| No | 0.88 | 9 | 1 | 1 | 29 | 75 | 5 | 1 | 7 | 56 | 21 |
| Yes | 0.12 | 2 | 1 | 0 | 4 | 13 | 1 | 0 | 1 | 13 | 4 |
| Do not know | 0.00 | 0 | 0 | 0 | 0 | 2 | 0 | 0 | 0 | 0 | 0 |
| Prefer not to answer | 0.00 | 0 | 0 | 0 | 0 | 2 | 0 | 1 | 0 | 1 | 0 |
| **Neuroticism score** | 4.12 (3.27) | 5.9 | 1.5 | 6.0 | 5.0 | 4.9 | 3.6 | 1.0 | 2.0 | 4.7 | 5.7 |
| Number of incorrect matches in round | 2.33 (3.12) | 0.4 | 2.0 |  | 0.8 | 0.7 | 0.5 |  | 0.6 | 0.6 | 0.7 |
| Mean time to correctly identify matches | 562.8 (117.6) | 523.2 | 588.0 | 477.0 | 588.8 | 581.1 | 505.3 | 538.5 | 617.7 | 592.6 | 602.3 |
| Fluid intelligence score | 6.16 (2.15) | 6.5 | 5.5 |  | 5.5 | 6.1 | 4.0 |  | 6.0 | 6.2 | 3.9 |

Although the numbers of subjects carrying PTVs in each gene are small and so no formal statistical tests were performed, the summary results shown in Table 1 do allow some conclusions to be drawn. Overall, there is little to suggest that PTVs in these genes cause substantial educational or occupational impairment. It may be worth noting that the proportion of people reporting having no qualifications in the whole UK Biobank sample is 0.09 whereas the numbers for subjects with PTVs in *TRIO* and *GRIN2A* are 8/32 and 4/7 respectively. However, carriers of PTVs in these or the other genes do not in general have obviously impaired occupational functioning. Carriers of PTVs in *HERC1* and *RB1CC1* have a somewhat reduced tendency to report excellent or good general health and are somewhat more likely to report having seen a GP, but not a psychiatrist, for problems related to anxiety or depression.

The detailed information for subjects bearing PTVs in all genes apart from *CACNAG1* was examined and findings are summarised below for each gene. Subjects not mentioned specifically appeared to be essentially normal from a neuropsychiatric point of view. The details provided in the summaries below are presented in a way aimed at maintaining the confidentiality of the information.

### *SETD1A* (11 subjects)

One subject had a diagnosis of schizophrenia, was taking an antipsychotic and was unable to work because of sickness or disability. One subject was taking an antidepressant and reported minor, persistent affective symptoms with no formal diagnosis and had seen a GP but not a psychiatrist for mood problems and also had diagnoses of fibromyalgia and menstrual problems.

### *CUL1* (2 subjects)

One subject denied a formal diagnosis of bipolar disorder or depression but reported having had antidepressant medication prescribed for more than two weeks and a number of features possibly related to a previous affective disorder including racing thoughts, prolonged, profound depression, thinking life not worth living and extreme irritability.

### *XPO7* (1 subject)

No suggestion of significant neuropsychiatric problems.

### *TRIO* (34 subjects)

Of seven subjects with no qualifications, one had suffered an episode of major depression and one reported being unable to work because of sickness or disability, but had hypertension, bone disorder and ulcers. Another four of these subjects had major recurrent depression, of whom one had seen a psychiatrist.

### *SP4* (6 subjects)

One had seen a GP with what appeared to be minor affective symptoms but no subjects appeared to have significant neuropsychiatric problems.

### *GRIA3* (2 subjects)

No suggestion of neuropsychiatric problems.

### *GRIN2A* (8 subjects)

One had no qualifications but was in paid employment. They had seen a GP and psychiatrist and had been assigned formal diagnoses of depression, anxiety, developmental disorder and epilepsy along with a number of physical health problems. They were taking an antidepressant and a sedative antipsychotic as well as medication for physical problems. Three others also had no qualifications but were all employed or retired and showed no suggestion of any neuropsychiatric problems.

### *HERC1* (70 subjects)

29 subjects appeared to have clinically significant depression and/or anxiety, of whom one had been diagnosed with an episode of psychotic depression and two others were possibly bipolar.

### *RB1CC1* (25 subjects)

One subject had been diagnosed with a disorder of scholastic skills, had no qualifications, a number of anxiety related diagnoses and had prolonged reaction time with low fluid IQ. Six other subjects appeared to have clinically significant anxiety and/or depression while another seven had milder affective symptoms.

In summary, one subject with a PTV in *SETD1A* had a diagnosis of schizophrenia. one with a PTV in *HERC1* had psychotic depression and two subjects seemed to have developmental disorders, one with a PTV in *GRIN2A* and one with a PTV in *RBCC1*. There seemed to be somewhat increased rates of affective disorders among carriers of PTVs in *HERC1* and *RB1CC1*. A number of subjects with PTVs in other genes also had affective disorders but not clearly at levels which were higher than expected. Aside from this, inspection of the individual clinical fields did not reveal that subjects with PTVs in these nine genes tended to have any sub-clinical features of schizophrenia. They were not prescribed antipsychotics and there were no problems noted which might suggest undiagnosed serious mental illness or general impairments in educational, occupational or social functioning.

## Discussion

The high prevalence of PTVs in *CACNA1G* casts doubt on the previous conclusion that such variants in this gene have a role in schizophrenia aetiology. This issue will be resolved as and when additional case-control samples are analysed and will not be considered further here.

With respect to the other nine genes, the overall impression that emerges from studying this sample of carriers of PTVs among UK Biobank participants is that the effect of PTVs in these genes tends to be dichotomous. While they may substantially impact the risk of developing schizophrenia, if this does not happen then most carriers appear to be phenotypically normal. They do not tend to be somewhat impaired or to suffer from low grade psychotic symptoms or otherwise to have a sub-clinical manifestation of major psychiatric illness. There is a suggestion that carriers of PTVs in *HERC1* and *RB1CC1* may be at increased risk of developing depression and anxiety disorders but this is a post hoc observation which would need to be specifically tested in new samples, for example when sequence data for the remaining UK Biobank participants becomes available.

The results obtained for PTVs in these genes contrast with those reported for CNVs impacting schizophrenia risk (Kendall *et al*., 2019). As stated above, carriers of these CNVs who do not have schizophrenia do tend to have impaired educational and occupational functioning and hence to some extent could be regarded as having a subclinical manifestation of at least some of the negative features of the disease. With the partial exception of the *NRXN1* deletion, all these CNVs impact multiple genes and hence could be regarded as potentially having a broader impact on development, some also being associated with physical abnormalities. Several are also associated with intellectual disability and overall they are more commonly found in subjects with schizophrenia who have below normal IQ (Lowther *et al*., 2017). In comparison, PTVs in *SETD1A* are associated with developmental disorder as well as schizophrenia but it not seem to be the case for PTVs in the other nine genes implicated by SCHEMA (Singh and on behalf of the Schizophrenia Exome Meta-Analysis (SCHEMA) Consortium, 2022). Instead, de novo missense variants rather than PTVs in *TRIO, CACNA1G* and *GRIN2A* were associated with developmental disorder. Thus it seems that PTVs in these genes seem to confer a relatively specific risk for schizophrenia.

The liability-threshold model is extensively used in statistical genetics, including in schizophrenia genetics. It postulates a hidden quantitative variable called the liability such that everybody whose liability exceeds a given threshold develops disease and everybody else does not. The liability is a mathematical fiction which allows one to apply statistical analyses suitable for quantitative variables to binary phenotypes. However this does not mean that such a quantitative variable truly exists. There is no hidden trait which we could potentially measure one day which would neatly divide cases from controls. Thus, while we may use schizophrenia liability in statistical analyses when this is convenient we must avoid the temptation to think that this implies that schizophrenia itself is a quantitative trait or that risk factors which have additive effects in a logistic regression model necessarily act to make people more or less severely affected. We have shown elsewhere that the distribution of psychotic traits in the general population and among subjects with schizophrenia is very much not compatible with the conception of schizophrenia as simply representing the extreme of a trait which is normally distributed in the population (Curtis and Derks, 2018). The current findings reinforce further that schizophrenia is an illness which, doubtful cases aside, one either does or does not have. PTVs in these genes have large effects on risk but carrying one does not make somebody “a little bit schizophrenic” just as smokers do not have “a little bit of lung cancer” - they have lung cancer or they do not. The notion that risk factors can have big effects on probability of developing disease rather than necessarily affecting severity of disease fits well with the medical model of illness. In this model, a number of genetic and environmental risk factors, when combined with processes which may be essentially stochastic, may result in a set of pathological changes which are associated with a characteristic set of signs and symptoms qualitatively different from a normal phenotype (Mitchell, 2018). The presence of a risk factor enhances the probability of these events occurring but may not itself have an overt effect on the phenotype if illness does not occur.

The PTVs implicated in the SCHEMA study have large ORs compared to common variants and if a patient with schizophrenia were found to carry such a PTV then it could legitimately be regarded as being causal. Nevertheless, if we assume a background lifetime prevalence for schizophrenia of 1% then for any PTV with OR less than 50 a carrier is more likely to be unaffected than affected. Most carriers of these PTVs will not develop schizophrenia and the present work seems to show that the overwhelming majority of these unaffected carriers do not suffer from subclinical levels of impairment but rather that in terms of neuropsychiatric symptoms they appear to be similar to the general population.

## Data Availability

The raw data is available on application to UK Biobank. Detailed results are not provided in order to protect confidentiality. Code and scripts use to perform the analyses are available at https://github.com/davenomiddlenamecurtis.

https://www.ukbiobank.ac.uk/

## Acknowledgments

This research has been conducted using the UK Biobank Resource, application number 51119. This work was carried out in part using resources provided by BBSRC equipment grant BB/R01356X/1. The author wishes to acknowledge the staff supporting the High Performance Computing Cluster, Computer Science Department, University College London. The author wishes to thank the participants who volunteered for the UK Biobank project.

## Availability of data

The raw data is available on application to UK Biobank. Detailed results are not provided in order to protect confidentiality. Code and scripts use to perform the analyses are available at *https://github.com/davenomiddlenamecurtis*.

Supplementary information is available at MP’s website.

---

**Supplementary Table 1.**
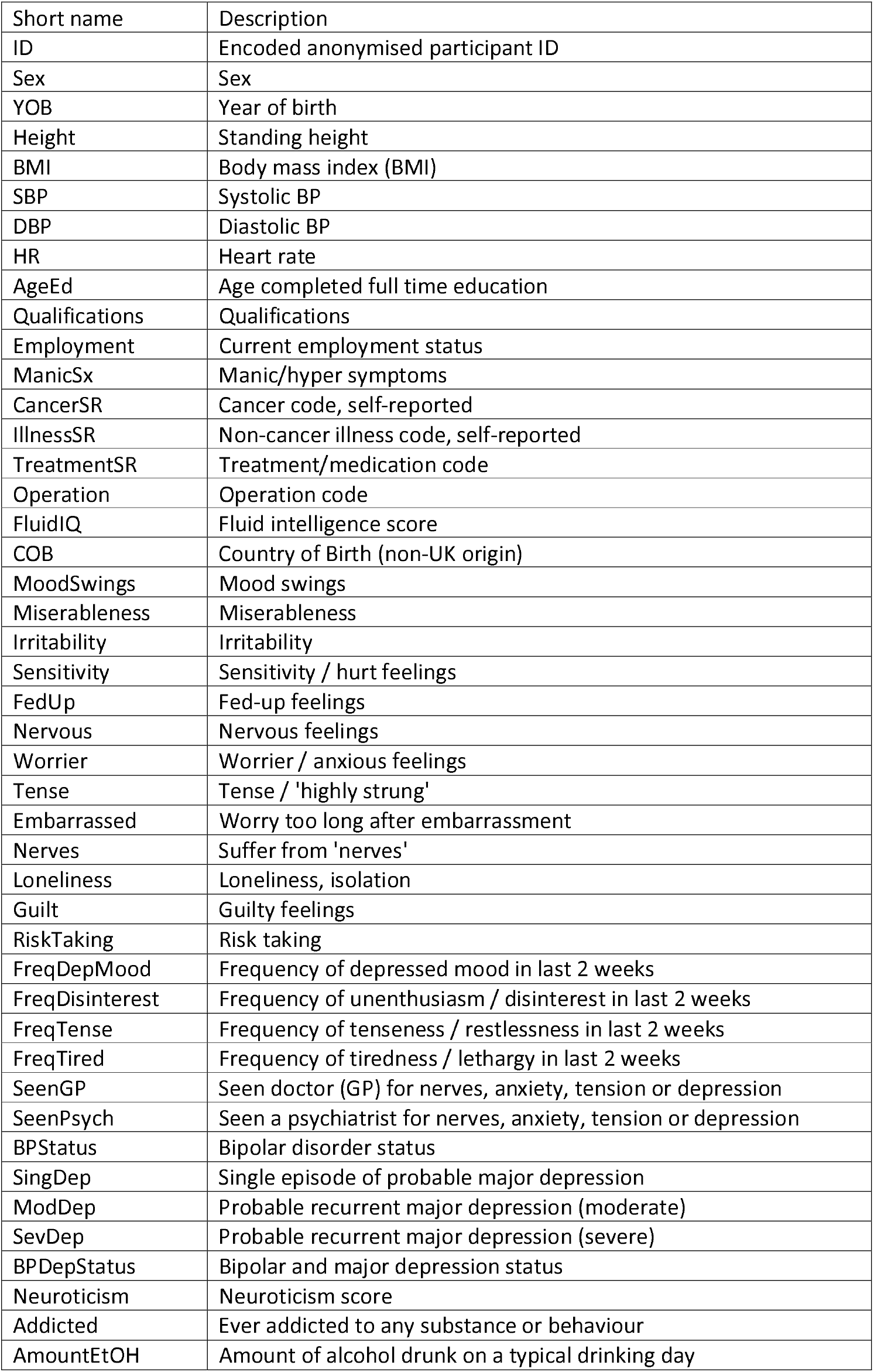
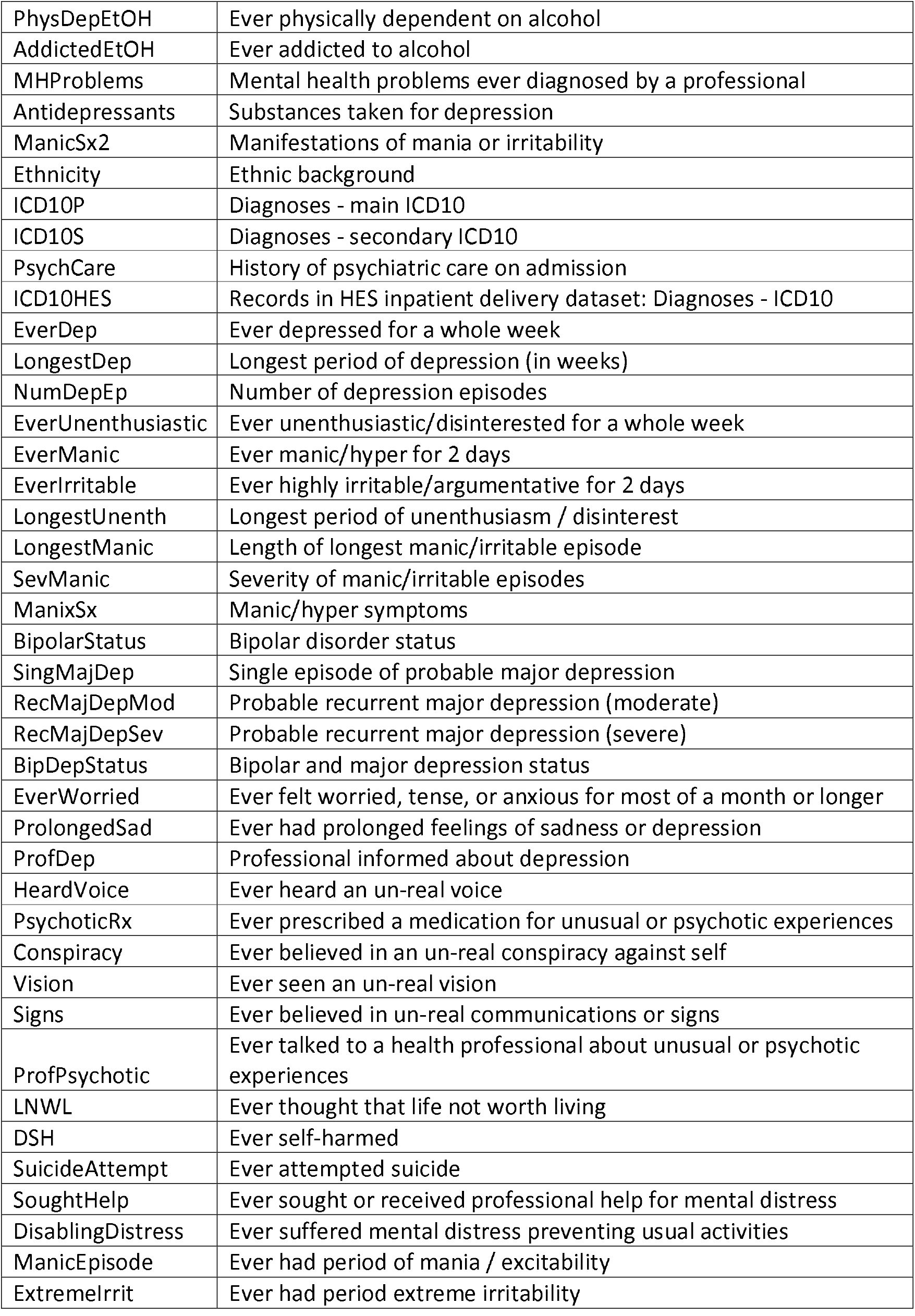
List of fields which were extracted to provide information about educational and occupational functioning and health information.

